# Do COVID-19 patients admitted to the ICU require anti-*Pneumocystis jirovecii* prophylaxis?

**DOI:** 10.1101/2020.05.18.20105296

**Authors:** Alexandre Alanio, Sebastian Voicu, Sarah Dellière, Bruno Mégarbane, Stéphane Bretagne

**Affiliations:** Laboratoire de Parasitologie-Mycologie AP-HP, Groupe Hospitalier Saint-Louis-Lariboisière-Fernand-Widal, Université de Paris, and Institut Pasteur, CNRS, Unité de Mycologie Moléculaire, Centre National de Référence Mycoses Invasives et Antifongiques, URA3012, Paris, France; Réanimation Médicale et Toxicologique, AP-HP, Groupe Hospitalier Saint-Louis-Lariboisière-Fernand-Widal, Université de Paris, INSERM UMRS1144, Paris, France

**Keywords:** COVID-19, Pneumocystosis, Pneumocystis jirovecii

## Abstract

We are currently facing a frightening increase in COVID-19 patients admitted to the ICU. Aiming at screening for secondary pneumonia, we collected the data of our first twelve ICU patients who underwent bronchoalveolar lavage (BAL). Surprisingly, four were detected with Pneumocystis jirovecii (Pj) DNA and RNA, resulting in Pj prevalence of 17%. Pj is a ubiquitous ascomycetes fungus that thrives at the surface of type-I pneumocytes, specifically in human alveoli, leading to pneumocystosis in immunocompromised patients. Interestingly, none of our patients was immunocompromised per se before admission, while all presented the recognized risk factors for life-threatening COVID-19 infection. Observing such high prevalence in COVID-infected patients was unexpected. Almost all patients developed ARDS and received high-dose steroids to prevent worsening, as suggested by reports from China. In Pj-positive patients requiring steroids, prophylaxis was given to avoid the risk of pneumocystosis and increased lung inflammation that may compromise the outcome.We are strongly convinced that testing deep lung specimens for Pj in severe COVID-19 patients should be recommended and Pj-positive patients treated with steroids, and given anti-Pj prophylaxis. This message is important, given the high mortality rate of COVID-19 patients in the ICU.

## To the Editor

Severe acute respiratory syndrome coronavirus 2 (SARS-CoV2) is spreading pandemically with more than 3,500,000 diagnosed cases and 250,000 fatalities reported on May 07^th^ 2020 (https://coronavirus.jhu.edu/map.html). Approximately 5-10% of coronavirus disease 2019 (COVID-19) patients may require intensive care unit (ICU) management including mechanical ventilation and 30% may develop secondary pneumonia without identified etiology (1). Hospital-acquired bacterial or fungal superinfections, as described in critically ill patients with Influenza virus, can be suspected (2). Since pneumocystosis is usually reported in patients with T-cell immunodepression (3), less attention has been paid to *Pneumocystis jirovecii* in non-immunocompromised ICU patients although it accounts for 7% of the co-infections reported in those admitted with Influenza (4). Interestingly, COVID-19 patients may develop acute respiratory distress syndrome (ARDS) requiring adjunctive steroids or other immunomodulatory therapies, a well-known susceptibility factor for developing pneumocystosis (4). Here, we investigated the prevalence of *P. jirovecii* acid nucleic detection in respiratory specimens sampled to identify co-infections in COVID-19 patients admitted to the ICU.

## Methods

All consecutive patients admitted to the ICU between March 15^th^ and April 15^th^ 2020 were enrolled. This study was part of the French COVID-19 cohort registry conducted by the REACTing consortium (REsearch and ACTion targeting emerging infectious diseases) and directed by INSERM (Institut national de la santé et de la recherche médicale) and ISARIC (International Severe Acute Respiratory and Emerging Infection Consortium). Our institutional ethics committee approved the study (IDRCB, 2020-A00256-33; CPP, 11-20 20.02.04.68737). When possible, signed informed consent was obtained from the patients or the next of kin.

Upon reception in the laboratory, all specimens including bronchoalveolar lavage (BAL) fluids and dTT-treated aspirations (dTT 1X at 37°C for 15min) were centrifuged, suspended in 200μL of water and submitted to extraction (whole nucleic acids extraction) using the GeneLead VIII extractor-thermocycler™ (Precision System Science, Matsudo, Japan). *P. jirovecii* reverse transcriptase quantitative PCR (RTqPCR) was performed to amplify mtSSU and mtLSU RNA and DNA of *P. jirovecii* using the new R-DiaPnJ kit™ (Diagenode, Liege, Belgium). β-D-glucan was tested using the Fungitell kit™ (Cape Cod Inc, Falmouth, US) as recommended by the manufacturer. Data are presented as median [range] or percentages as appropriate. Comparisons were performed using Mann-Whitney or exact Fisher tests as required. *P*<0.05 was considered as significant.

## Results

Twenty-nine successive COVID-19 patients (M/F sex ratio, 2.6; age, 60 years [33-80]) with the usual risk factors for severe COVID-19 presentation were included (Table1). All patients except one were intubated on admission. Twenty-seven patients (93%) who developed ARDS received corticosteroids once mechanically ventilated. Respiratory samples included 27 BALs (93%) and two bronchial aspiration fluids (7%). In 5/29 patients (17%), *P. jirovecii* RTqPCR was positive. The median quantitative cycle value was 32.4 [28.9-36.5].

**Table 1.** Characteristics of twenty-nine critically ill COVID-19 patients according to *Pneumocystis jirovecii* detection in the respiratory samples

| Parameters | *P. jirovecii* RTqPCR-positive patients<br>(n=5) | *P. jirovecii* RTqPCR-negative patients<br>(n=24) | *P Value* |
| --- | --- | --- | --- |
| Age (years) | 59 [41-79] | 60 [33-80] | 0.64 |
| Gender (M/F) | 1.5 | 3 | 0.60 |
| Diabetes, N (%) | 1 (20) | 10 (42) | 0.62 |
| Hypertension, N (%) | 3 (60) | 11 (46) | 0.65 |
| Obesity, N (%) | 0 | 7 (29) | 0.30 |
| Ischemic heart disease, N (%) | 1 (20) | 5 (21) | 0.99 |
| Prior steroid therapy, N (%) | 2 (40) | 2 (8) | 0.13 |
| PaO<sub>2</sub>/FiO<sub>2</sub> (mmHg) | 167 [79-290] | 162.5 [42-480] | 0.59 |
| Time from symptom start to SARS-CoV2 detection<br>(days) | 7 [4-13] | 7.5 [3-30] | 0.92 |
| Time from symptom start to tracheal intubation<br>(days) | 8 [5-16] | 8 [3-33] | 0.72 |
| Time from tracheal intubation to BAL (days) | 1 [0-5] | 3 [0-20] | 0.25 |
| Lymphocyte count (G/L) | 0.8 [0.3-1.5] | 1.1 [0.3-2.8] | 0.99 |
| BAL macrophages, N (%) | 53 [49-72] | 26 [4-46] | **0.0003** |
| BAL neutrophils, N (%) | 23.5 [10-32] | 46 [3-87] | 0.29 |
| BAL lymphocytes, N (%) | 18 [10-39] | 23 [1-74] | 0.83 |
| β-D-glucan (pg/mL) (N=13) | 16.3 [7.0-105.0] | 13.1 [7.8-523.0] | 0.60 |
| Viral load, (cycle) | 23 [17-32] | 28 [15-37] | 0.34 |
| Steroid administration in the ICU, %† | 4 (80) | 17 (71) | 0.99 |
| Outcome (death / in ICU / discharged), N (%) | 1 (20) / 1 (20) / 3 (60) | 9 (38) / 3 (12) / 12 (50) | 0.1 |

BAL, bronchoalveolar lavage; ICU, intensive care unit; RTqPCR, reverse transcriptase quantitative polymerase chain reaction;
†Steroid regimen, dexamethasone intravenous dose of 20 mg once daily from day 1 to day 5, followed by 10 mg once daily from day 6 to day 10.

No significant differences were observed between *P. jirovecii* RTqPCR-positive and negative patients in terms of age, risk factors for severe COVID-19 including prior steroid therapy, severity of ARDS, viral load, blood lymphocyte count, time between intubation or onset of symptoms and SARS-CoV2 detection and time between onset of symptoms, intubation or SARS-CoV2 detection and respiratory specimen sampling. Interestingly, the proportion of macrophages in BAL fluids was significantly higher in *P. jirovecii* RTqPCR-positive *versus* negative patients (*P*=0.003; Figure 1), by contrast to the proportions of neutrophils and lymphocytes.

**Figure 1.** Distribution of the proportion of macrophages in the bronchoalveloar lavage (BAL) with positive or negative *Pneumocystis jirovecii* reverse transcriptase quantitative PCR (PjRTqPCR)

Three out of five *P. jirovecii* RTqPCR-positive patients received co-trimoxazole as prophylaxis. To date, one patient has died, one patient is still mechanically ventilated and the three others including the two patients who were not treated with co-trimoxazole have been discharged from the hospital.

## Discussion

We found an unexpectedly high proportion of critically ill COVID-19 patients detected with *P. jirovecii* (17%) as compared to previous findings in influenza patients (~7%) (4).

The presence of *P. jirovecii* in the healthy adult population has been measured using oropharyngeal wash samples obtained by gargling and examined by conventional or nested PCR methods.^5^ However, experts agree that the reported prevalence (~20%) has been overestimated due to technical issues such as contamination with amplicons responsible for false positives (3). In our experience using qPCR, around 15% of BAL fluids are positive in immunocompromised patients (3), supporting the proposal that the prevalence we observed in the apparently non-immunocompromised COVID-19 patients is actually elevated. These patients mostly exhibit marked lymphopenia and alterations in lymphocyte functions (6), likely explaining the high-rate of *P. jirovecii* detection. Nevertheless, β-D-glucan concentrations measured in the respiratory fluid specimens obtained in our five *P. jirovecii* RTqPCR-positive patients were lower than 120pg/mL supporting limited fungal loads in the lung alveoli (7). Of note, in two out of our nine tested *P. jirovecii* RTqPCR-negative patients, higher β-D-glucan concentrations (450 and 523pg/ml) lead to the diagnosis of putative invasive aspergillosis, another fungal infection of risk in COVID-19 patients (8).

Surprisingly, the BAL macrophage proportion was particularly elevated in our *P. jirovecii* RTqPCR-positive COVID-19 patients. In immunocompromised adults with pneumocystosis, BAL macrophage proportions of ~45% have been reported (9). Interestingly, in non-HIV patients, the count and proportion of BAL macrophages was significantly increased in mild compared to severe pneumocystosis, i.e. 40.7% versus 28.3% and 76 *versus* 50cells/μl, respectively (10). Taken together, these findings strengthen the hypothesis that *P. jirovecii* presence in the deep lung may influence local immunity and enhance inflammation.

Here, three out of five *P. jirovecii* RTqPCR-positive patients received co-trimoxazole as prophylactic regimen, based on the treating physician’s decision. Whether a positive result should be an indication to consider administering co-trimoxazole, at least at prophylactic dosage in COVID-19 patients remains questionable.

Our study limitations include the relatively small number of patients, the single-center setting, and the short study period. However, to the best of our knowledge, this is the first study evaluating the prevalence of *P. jirovecii* in COVID-19 patients. Because we focused on critically ill COVID-19 patients, prevalence of *P. jirovecii* in less severe patients remains to be determined.

In conclusion, based on our findings, we advocate systematically searching for *P. jirovecii* in deep respiratory specimens in critically ill COVID-19 patients. We believe that this strategy may be useful in limiting enhanced inflammation due to the presence of *P. jirovecii* in the lung and avoiding inter-patient *P. jirovecii* transmission.

## Data Availability

Data will be foully available for the community

## Author Contributions

*Concept and design:* Alanio, Mégarbane, Bretagne

*Patient care:* Voicu, Mégarbane.

*Microbiological analysis:* Alanio, Dellière, Bretagne

*Acquisition, analysis, or interpretation of data:* All authors.

*Drafting of the manuscript:* Alanio, Mégarbane, Bretagne

*Critical revision of the manuscript for important intellectual content:* All authors.

Dr Alanio had full access to all of the data in the study and takes responsibility for the integrity of the data and the accuracy of the data analysis.

## Acknowledgment

The authors would like to thank Mrs. Alison Good (Scotland, UK) for her helpful review of the manuscript. We would also like to thank our team who managed all COVID-19 patients and our lab technical staff who performed pneumocystis RTqPCR on COVID-19 positive specimens.

